# Mapping the Global Research and Clinical Trials in COVID-19

**DOI:** 10.1101/2020.06.27.20141788

**Authors:** Ranjana Aggarwal, Sujit Bhattacharya, Shubham Singh

## Abstract

COVID-19 has created an unprecedented level of research and innovation activity globally to bring out drug to control or cure the disease, and develop vaccine for long time prevention. A ‘new/better normal’ is emerging that is trying to push this time period for drug development and vaccine within a year and earlier than that from typical 8 to 10 years for drugs and 10 to 15 years for vaccine. This is happening due to multiple factors: strong policy push by different government that includes dedicated investment, speeding up regulation process, multiple agencies involvement, and multilateral bodies led by WHO trying to create a global platform, huge grants from private funding bodies, strategic linkages across the whole research and innovation value chain between firms, academic, research organisations and start-ups.

The paper maps the research papers and ongoing clinical trials to provide an informed view of the current status of the research and drug development activity as seen through the lens of research papers and clinical trials. The intended goal of this study is to help the research community and policy makers to keep track of highly relevant research and drug development in COVID-19.

## Introduction

The intended goal of this study is to help the research community and policy makers to keep track of highly relevant research and drug development (as seen through the lens of clinical trials) in COVID-19. Since the pandemic began, there were large number of studies trying to understand the structure of the virus, transmission and the mitigation measures. The studies helped to identify the key facts of the virus, initial response for containing the virus, efficacy of measures such as face masks, hand hygiene, to reduce the reproduction number of the virus etc. [1] [2] The focus has now shifted to clinical research and various organisations are collaborating to develop a cure for the virus or reduce the mortality rate.

Many clinical trials are centered on repurposed drugs. This repurposing strategy “old drugs, new use” is emerging as the most preferred strategy to bring a drug quickly to the market for treatment of this disease or address specific conditions that is emerging due to it. The rationale behind this strategy i.e. repurposing strategy is driven by sound logic. Drug development is a long process that covers clinical trials at different phase from preclinical (animal testing for efficacy and toxicity) to clinical trials at different phases (I to III). It includes testing of drugs on healthy persons for safety (phase I), on patients to assess efficacy (Phase II), further large scale testing to assess efficacy, effectiveness and safety (Phase III). The chances of failures are high at different stages which sinks the cost that was incurred. Keeping all these factors in consideration, it is estimated that a new drug to reach the market costs $800 million to $1000 million. [3] As a recent study in Lancet [4] cautions “Clinical trial literature is riddled with drugs that looked promising in small trials only to prove ineffective in bigger, more rigorous studies.” The present pandemic calls for quick interventions. Thus due to urgent need to bring a drug to the market, and the uncertainties involved, repurposed drug becomes a much safer pathway. Availability of the safety data along with addressing similar conditions and broad spectrum is available for these drugs as they are tested previously on other diseases and hence their clinical trials testing is accelerated. [5] Clinical trials thus can be much shorter and more directed and can have combination of phases for repurposed drugs. The clinical trials can concentrate on Phase II onwards. Thus repurposed drugs can be much faster to bring to the market. Thus it is not surprising to see this as the most common strategy adopted by research institutions/universities and firms for Covid-19.

Many repurposed drugs that are part of clinical trials can be distinguished by promising results for earlier coronavirus diseases (SARS-CoV-1 or MERS) or drugs that were effective antivirals. Solidarity trials undertaken by the WHO is also a repositioning drug strategy conducted on four drugs Remdesivir, Lopinavir/Ritonavir, Lopinavir/Ritonavir with interferon beta-1a. Clinical trials of these drugs are being undertaken in multiple countries that have given consent to this approach for treatments. WHO claims that thisglobal coordinated clinical trials will reduce the time taken by randomised clinical trials by 80%. Many drugs that were not so effective earlier in terms of success for the condition for which it was developed are also being revisited for this disease. *This is another type of repurposing*. A good example of this is the Gilead Sciences drug Remdesivir which is found to inhibit the activity of RNA-dependent RNA polymerase (RdRp)2 of SARS-CoV-2. [6] The drug according to Scavone et al. [7] “was initially developed as a treatment for Ebola and Marburg infections but did not demonstrate clinical efficacy” now is used in the COVID-19 treatment. National Institute of Allergy and Infectious Diseases (NIAID) is also conducting clinical trials on Remdesivir which is a multicentre, randomised, placebo controlled Phase 3 trial.

Other methods like convalescent plasma therapy and antibodies treatment are also showing promising results. Convalescent plasma is seen to be a prominent intervention in the clinical trials. It is a passive antibody therapy in which blood from the recovered patients is taken and transfused in a COVID-19 patient. The treatment was advised previously by WHO in treatment of Ebola and MERS diseases. [8] Studies that support this line of treatment for COVID-19 include the study published in Lancet [9] which states “The viral load [on patients] after convalescent plasma treatment was significantly lower on days 3, 5, and 7 after intensive care unit admission”. The study also pointed out the reduction in risk of mortality and suppression of viramia (presence of virus in the bloodstream). All the convalescent plasma trials are in advanced stages of clinical trials owing to quick approvals of this treatment as no severe adverse effects are seen to have been associated with this treatment. Elliy Lilly has developed an antibody LY-CoV555 which targets SARS-CoV-2 spike protein. Using plasma from recovered patients to treat others who have developed this disease is another active line of treatment. According to Gul et al. [10] “Convalescent blood products are the most promising potential treatment for use in COVID-19”.

Another line of research is for developing a quick and effective vaccine. For this various vaccine models are used like: weakened or inactivated virus model, DNA or RNA vaccines, viral vector vaccines, or vaccines based on protein subunits (spike proteins etc.) or virus like particles (empty virus shells without genetic material). An interesting insight of the vaccine development landscape can be seen from the WHO’s “DRAFT landscape of COVID-19 candidate vaccines” [11] a total of 133 vaccines candidates are in development around the world. Ten of these are in clinical evaluation and 123 in Pre-clinical evaluation.

The paper focusses on global research and drug development. A successful vaccine is most effective for prevention of this disease and it can prevent this type of pandemic we are seeing. The paper thus has this limitation as it does not capture the ongoing clinical trials for vaccine development of this disease.

### Objective and Research Questions

The study attempts to capture the influential research and insights of ongoing clinical trials surrounding COVID-19. It attempts to address the following research questions:

- To capture the key research papers and their characterisation?
- To draw insights from ongoing clinical trials?

Based on the above, we envisaged to draw policy implications for research and innovation surrounding this pandemic.

## Methodology

The data for this study was drawn from Dimensions database (www.dimensions.ai), an integrated linked database that provides data of funding agencies (grant), research publications, patents, clinical trials and key policy documents. The database captures clinical trials database from a number of international clinical trials registry and contains 557322 clinical trials data.

The COVID-19 research papers and clinical trials were extracted from Dimensions database using the search string “Covid-19” Or “SARS-CoV-2” Or “SARS-CoV2” Or “2019-nCoV” on 2 June, 2020 from this database in the full text. Global Research papers and clinical trials trends were also collected from 22 May 2020 to 2 June 2020 using the above search string. Altmetrics or article level metrics was chosen as the popularity qualifier.

Field of research (FOR) classification available on dimensions database was chosen as the research paper classifier. This diverse nature of FOR classification system unlike other systems available in dimensions like RCDC which is completely biomedical provided the basis for its selection. The 15 most frequently occurring FOR categories in the COVID-19 papers were identified from theabove168 classifications available. The papers under these categories were mapped to show some insights of the dispersion of the papers across key research themes. A paper could be placed in more than one category and thus summing up the counting of papers across categories would be considerably more than the total papers.

Through altmetrics score Top 10 most popular papers were identified. Data for twitter, news, facebook and blog mentions was collected from altmetrics.com. Audience and country where the paper is most popular was based on the type of tweeter (public, scientists, practitioners).

A total of 3428 Clinical trials were extracted on 2 June 2020. Co-occurrence map of 42 interventions occurring atleast 4 times in the clinical trials was constructed using text analytics software VOSviewer and Pajek. The co-occurrence calculates the number of times two or more words (concepts/topics) occur together. The paper has used the co-occurrence map to analyse the relationship between various RCDC categories and interventions.

## Results

A very intensive research activity and clinical trials can be observed surrounding COVID-19. There were 47052 research papers (39,955 published articles and 7097 preprints), and 3581 clinical trials covering various aspects of this disease as on 8 June, 2020. Figure 1 highlights the research papers published and clinical trials during the period from 22 May to 8 June, 2020.

**Figure 1:**
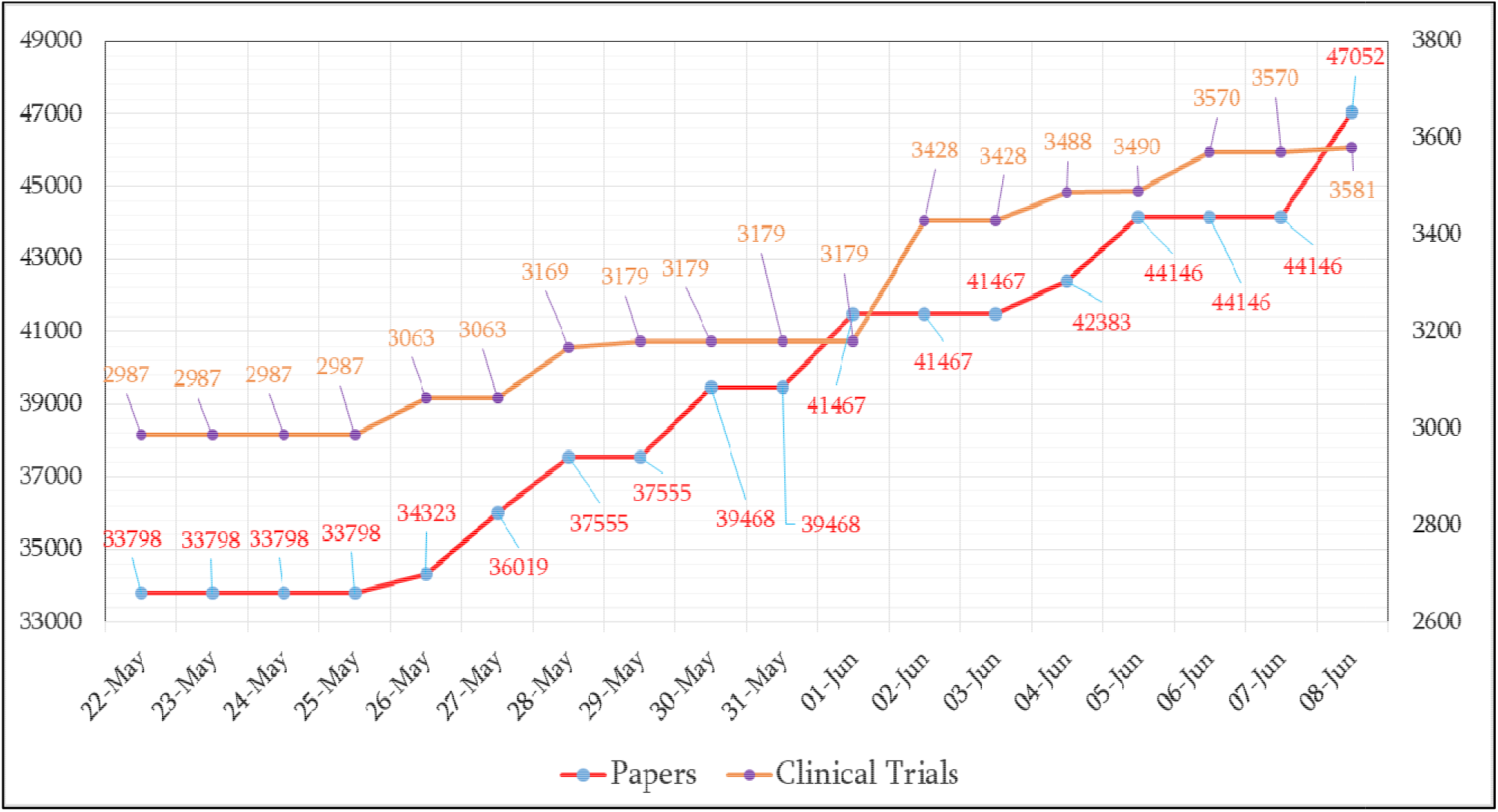
Global Papers and Clinical Trials Trend.

High quality scientific research is an essential pre-requisite for drug/vaccine development. Knowing the various characteristic of the SARS-COV-2 virus and the disease COVID-19 caused by it, the genome structure and phylogenetic analysis of the virus, methods of transmission, its various effects on the human body, treatment (efficacy, toxicology) etc. surrounds the drug/vaccine development. The intense science driven process behind drug/vaccine development is the primary reason behind the intensity and large volume of research. Fast tracking of COVID-19 research by various journals and quick availability on pre-print servers has also played a major role in this increase.

It was found that as many as 168 FOR classification category covered the research papers; from ‘medical and health sciences’, ‘clinical sciences’, to economics. This shows the wide impact of this disease as it in a sense motivates research activity across so many diverse domains (disciplines/sub-disciplines). Research papers are an indication of this dispersion of research activity. Figure 2 highlights the main domains wherein research papers are visible.

**Figure 2:**
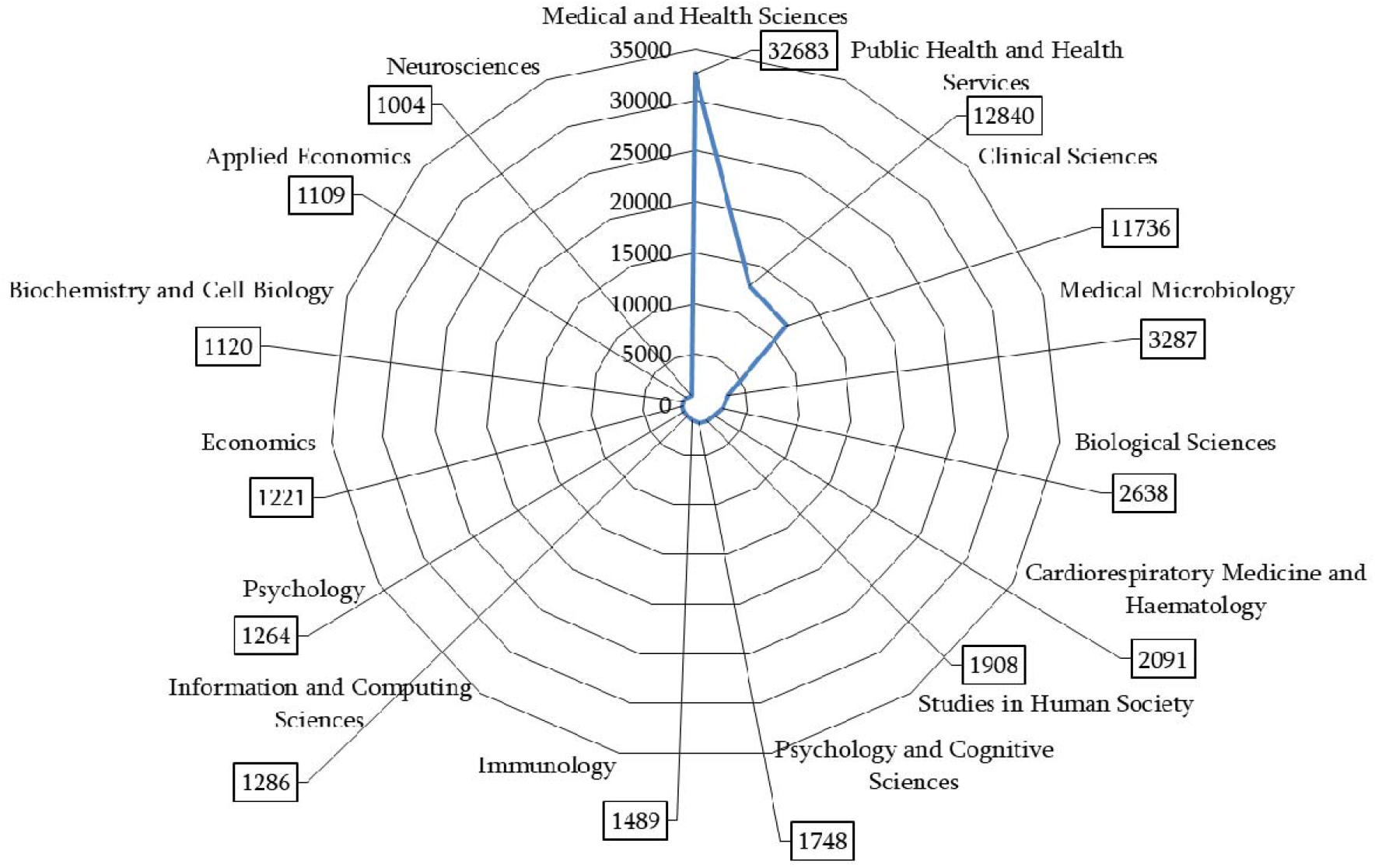
Main domains of COVID-19 research.

The above figure underscores again the wide diversity in terms of research topics being covered. The papers fall into more than one topic so the total count is much higher than the overall papers. The distribution topic wise shows along with research in various medical aspects of the disease, there is also intense research in economic and social aspects of it.

A further examination of the ten most popular papers (Table 1) and their content analysis provide deeper insight into this disease.

**Table 1:**
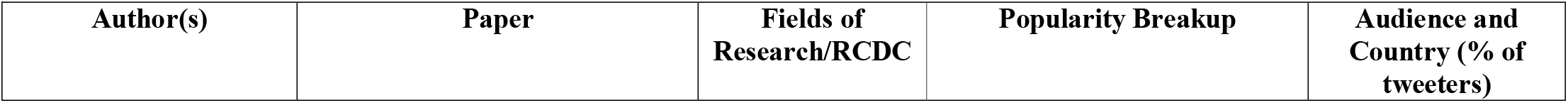
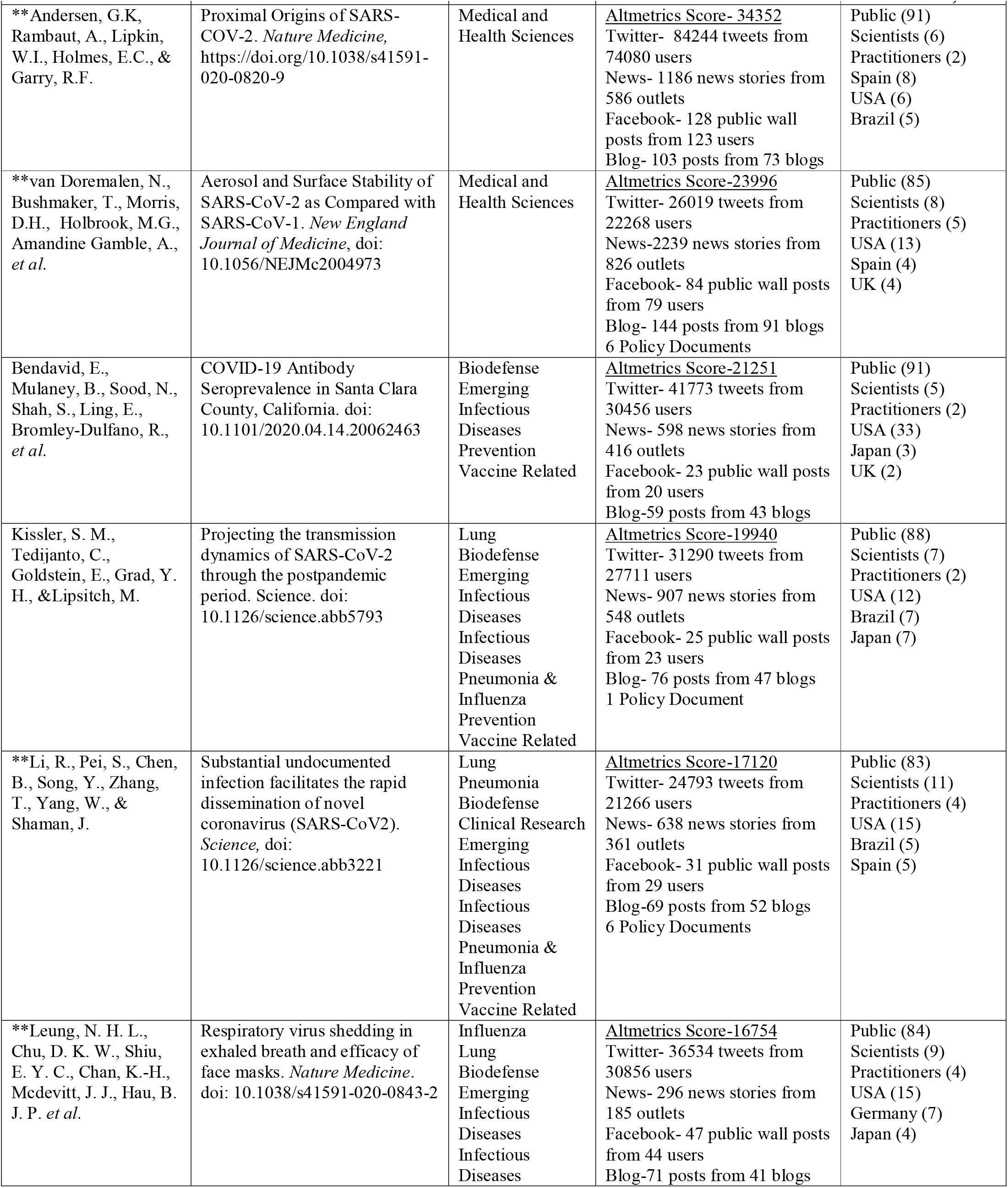
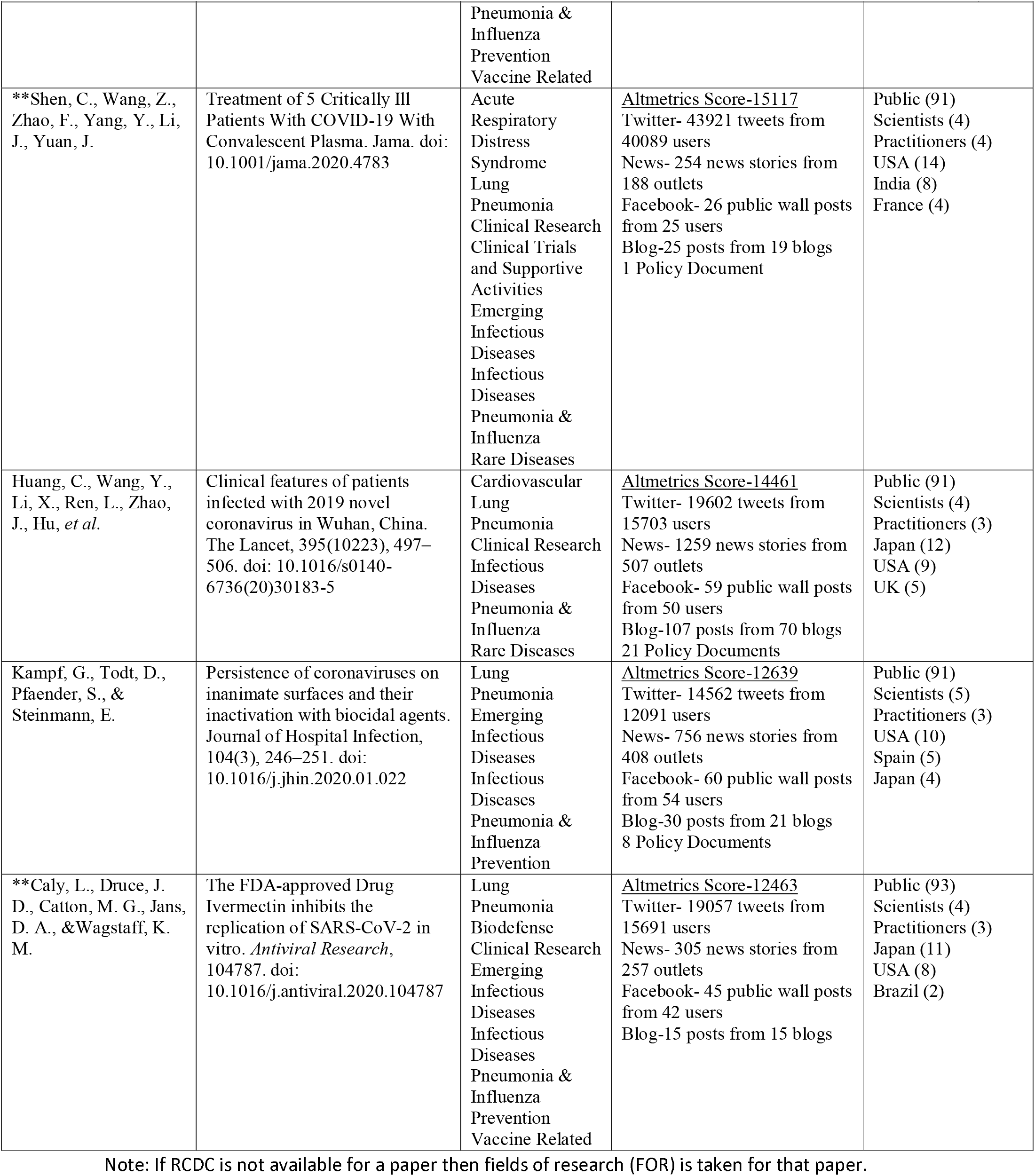
Highlighting Key Characteristics of the Ten Influential Papers.

The content analysis of the papers underscores in a sense what has been the influential aspect of the paper that has distinguished it from the large repository of papers surrounding this disease. It is also interesting to note that of the 10 popular papers as many as 6 papers have remained in the top ten papers over a period of time (these papers indicated by start mark in the Table) This further demonstrates their impact.

### Content Analysis of the above Ten Papers

Anderson *et al. (2020)*, the study most popular on social media platforms (number of tweets more than three times the next popular paper) showed that “SARS-COV-2” is the seventh coronavirus to infect humans”. On the basis of comparative analysis of genomic features of SARS-CoV-2 with other alpha and beta coronaviruses the authors claimed that this virus is optimized for binding to the human receptor ACE2 and the spike proteins of this virus has a polybasic (furin) cleavage site at S1-S2 boundary which may be important to determine its impact on transmissibility and pathogenesis in animal models. It further claimed that SARS-CoV-2 is not a product of purposeful manipulation, most likely the result of natural selection of human or human-like ACE2 receptor. *The different aspects covered by this disease can be seen highly important and thus contributed to its online impact*.

van Doremelen *et al. (2020)* analysed the “aerosol and surface stability of SARS-COV-2 and compared it with SARS-COV-1, the most closely related human coronavirus. The study highlighted the need to protect from five environmental conditions: aerosols, plastic, stainless steel, copper and cardboard. Further it showed that SARS-COV-2 remains viable in aerosols for 3 hours, on plastic and stainless steel for 3 days, 4 hours on copper and on cardboard for 24 hours. The study found that the stability of SARS-COV-1 is similar on plastic, stainless steel and aerosols to SARS-COV-2 and different to SARS-COV-2 on cardboard (8 hours) and copper (8 hours). It was also indicated that though the stability of both viruses is similar, both show exponential decay in virus titer, the difference in the epidemiologic characteristics of these viruses may arise due to high viral loads in the upper respiratory tract and the potential for infected persons to shed and transmit the virus while asymptomatic. *The implications of this study can be clearly seen in the preventive measures of COVID-19. This study was also cited in policy documents*.

Leung *et al*. (2020) explored “the importance of respiratory droplet and aerosol route of transmission” by quantifying the “amount of respiratory virus namely coronavirus, influenza virus and rhinoivirus in exhaled breath of participants” that have acute respiratory virus illness (ARI). The 246 participants were divided in two groups, one wearing surgical face mask and other not wearing face mask. The study found that surgical face masks can efficaciously reduce the respiratory droplet emission of influenza virus particles but not in aerosols. They also found that surgical face masks can be used by ill patients of COVID-19 to reduce “onward transmission”. Face mask is getting increasing attention and now being incorporated as essential guideline in health policies of different countries. *The paper provided a good empirical support to this i*.*e. face masks*.

Li *et al. (2020)* estimated that 86% of COVID-19 cases went undocumented in China prior to their travel restrictions. The study also estimated that the undocumented cases contagiousness or transmission rate was 55% of documented infections, yet 79% of documented infection cases were due to these undocumented infections. The suggestion of this study that undocumented infections “isolation and identification is necessary to fully control the virus” is very important and the spread of this virus may be seen as a consequence of this. This study also was cited in policy documents.

Caly *et al*. (2020) reported that Ivermectin, an FDA approved broad spectrum parasitic agent previously shown to have anti-viral activity against a broad range of viruses in vitro, inhibits the SARS-COV-2. The study found that a single dose of Ivermectin added to Vero-hSLAM cells 2-hours post infection with SARS-CoV-2 isolate Australia/VI01/2020 2 was “able to reduce the viral RNA by approximately 5000-fold within 24-48 hours”. They also hypothesized the mode of action through inhibiting IMPα/β1-mediated nuclear import of viral proteins. Doctors are struggling to control this dangerous disease. *A study like this which gives some hope is quickly tracked which is indicated by its high altmetrics score*.

Shen *et al*. (2020) study examined convalescent plasma, containing neutralizing antibody, transfusion benefit in treatment of critically ill COVID-19 patients. The clinical trial was conducted on 5 critically ill patients with COVID-19 and acute respiratory distress syndrome (ARDS) along with certain other conditions. The study found decline in viral load, clinical conditions of patients improved as “indicated by body temperature reduction, improved PAO_2_/FIO_2_ and chest imaging”. Though the sample size is limited and requires further clinical trials for potential effectiveness but this treatment is now being incorporated in many countries.

Bendavid *et al*. reported a new dimension to address epidemic models, projections and public policies on COVID-19 on the basis of measurement of seroprevalence of antibodies to SARS-CoV-2 in Santa Clara County. Using lateral flow immunoassay, a World Health Organization protocol for COVID-19 antibody testing, serological testing was conducted for SARS-CoV-2 antibodies in a sample of 3300 people selected on the basis of three data elements: zip code of residence, sex and ethnicity/race. On the basis of seroprevalence data of antibodies to SARS-CoV-2 (between 2.49% to 4.16%) among the population, they estimated that the number of infections is much higher of the order of 50-85 fold in Santa Clara than indicated by the number of confirmed cases and the fatality rate is 0.12-0.2% which is much lower than the reported average increase of 6% daily as of April 10, 2020. This well-measured data on population prevalence estimates provided important understanding to calibrate epidemic stage, calculate fatality rates and frame public policy decisions. The paper is attracting attention of several research groups worldwide who are testing population samples for SARS-CoV-2 antibodies and citing this paper frequently.

Kampf *et al*. provided a comprehensive review on the persistence of human and veterinary coronaviruses on different type of inanimate surfaces as well as their inactivation strategies with biocidal agents used in surface disinfectants. The data showed that human coronaviruses such as Severe Acute Respiratory Syndrome (SARS-CoV), Middle East Respiratory Syndrome (MERS-CoV) and endemic Human coronavirus strain (HCoV-229E) can remain infectious on inanimate surfaces like steel, aluminium, metal, wood, paper, glass, plastic, PVC, silicon rubber, latex, disposable gown, ceramic and Teflon for from 2 hours to 9 days. It also captured the effect of temperature, humidity and concentration of inocculum (viral titter) on the persistence of coronavirus. Inactivation of these coronaviruses by a variety of commonly used biocidal agents namely ethanol (78-95%), 2-propanol (70-100%), combination of 1-and 2-propanols (45% and 30%), glutardialdehyde (0.5-2.5%), benzalkonium chloride (0.05-0.2%), chlorohexidine digluconate (0.02%), sodium hypochlorite (0.001-0.21%), hydrogen peroxide (0.5%), formaldehyde (0.009-1%) and povidone Iodine (0.23-4%) has also been compiled. The study indicated that the most efficient disinfectants to inactivate these viruses on the surfaces within 1 minute are 62-71% ethanol, 0.5% hydrogen peroxide or 0.1% sodium hypochlorite and are also expected to disinfect SARS Cov-2 similarly. As no antiviral treatment for SARS-CoV-2 has been proven to be effective till date, contamination and prevention of further spread is the key strategy to stop the ongoing outbreak. This review article gained popularity as it provided inputs about the best disinfectants to be used to ensure disinfection of frequently touched surfaces, hand hygiene in healthcare set ups and for public usages at large to combat the spread of pandemic. *Huang et al*. reported the epidemiological, clinical, laboratory and radiological features of patients infected with 2019 novel coronavirus in Wuhan, China. Data of 41 laboratory-confirmed patients for the period Dec.16, 2019 to Jan.2,2020, collected from clinical charts, nursing records, laboratory findings and chest x-rays revealed similarities of clinical features between 2019-nCoV and previous betacoronaviruses such as SARS-CoV and MERS-CoV. Some of the important features highlighted in this article about the illness were: most of the infected patients were men (73%) with less than half having co-morbidities like diabetes, hypertension, cardiovascular disease; 66% patients had direct exposure to Huanan seafood market; most patients suffered with fever, dry cough, dyspnoea, fatigue and bilateral ground-glass opacities on chest CT scans and only a few with haemoptysis and diarrhoea. The pathophysiological studies of patients infected with 2019-nCoV indicated that amount of proinflammatory cytokines IL1B, IFN[_, IP10 and MCP1 and TNFα, responsible for pulmonary inflammation and extensive lung damage, were increased in serum. Patients with higher concentrations of GCSF, IP10, MCP1, MIP1A and TNFα required ICU admission suggesting that cytokine storm was associated with disease severity. This article gained significance as it was one the earliest reports to provide evidence based information on common complications associated with infection with COVID 19 and the line of treatment adopted by the doctors in Wuhan.

Kissler*et al*. projected SARS-CoV-2 transmission model for the pandemic period using time series data from the USA for betacoronaviruses namely OC43 and HKU1.The assessments were based on role of seasonal variation, duration of immunity and cross immunity on the transmissibility of these two viruses. Through model simulations they predicted some patterns of outbreaks: substantial outbreaks in autumn/winter than winter/spring; annual outbreaks if SARS-CoV-2 establishes short-term immunity on the order of 40 weeks while biennial for long term immunity of two years; seasonal variation in SARS-CoV-2 differ between geographical locations as for influenza; decline or elimination of incidences of betacoronaviruses if SARS-CoV-2 induces 70% cross immunity against HCoV-43 and HCoV-HKU1; elimination of transmission for upto three years if low level of cross immunity (30%) is induced after the initial, most severe pandemic wave. Complimentary interventions such as expanding critical care demand and effective therapeutics along with success of social distancing would hasten the acquisition of herd immunity. The study emphasized the urgency of longitudinal serological studies to be undertaken to determine the extent and duration of immunity of SARS-CoV-2 and epidemiological surveillance to be maintained in the coming years to anticipate the possibility of resurgence. The study is important as it gives valuable inputs for the prediction of outbreaks so that world is better prepared in terms of healthcare, therapeutics and economic front to deal with it effectively.

As the content analysis highlights, each of the paper covers various aspects of the disease, from understanding the structure of the virus, effect of the disease (complications, conditions), the way it spreads and interventions that can be effective to contain its spread, drugs/treatments that show initial promise. The research papers have influenced the drug development process as they have provided initial evidence/pathway. They have also influenced government intervention measures to prevent the spread and contain the disease. Some evidence of this also comes from the various Altmetric measures.

### Clinical Trials

As on June 8, 2020 there are 3581clinical trials on Covid-19. However, for some details were not available. During this period there were 1659intervention based clinical trials and 1727 observational clinical trials, 129 retrospective studies, 5 new treatment measure clinical study. Figure 3 shows the key 42 interventions occurring in at least 4 clinical trials.

**Figure 3:**
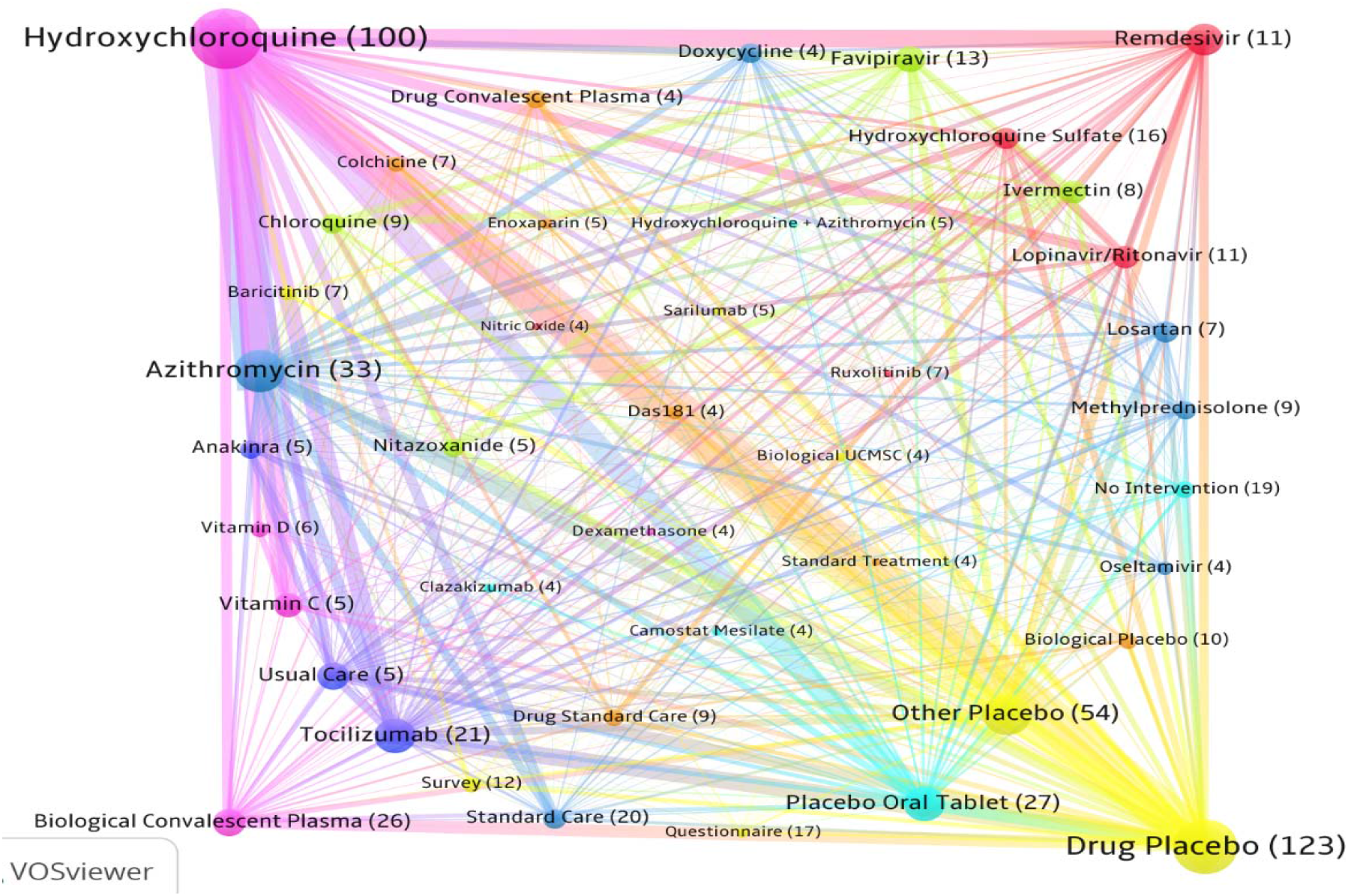
Co-occurrence map of Interventions in Global Clinical Trials. Note: The values in brackets show the frequency of these interventions. The thickness of the linkages shows the strength with which two interventions are linked together.

Deeper analysis based on of the linkages as shown by the map draws attention to some key insights of the ongoing clinical research. One can observe Hydroxychloroquine, Azithromycin, Remdesivir, and Tozilizumab prominently visible in the map. All these drugs are repurposed drugs. The clinical trials are being undertaken to find out their efficacy for the treatment of Covid-19. The drugs (Remedesvir, Lopinavir/Ritonavir, Chloroquine and Hydroxychoroquine) both currently and previously included in the WHO solidarity trials can be seen in the co-occurrence map.

Strong linkage of these drug trials to Placebo underscores that trails are not ‘open labels’ wherein participants know the treatment they are getting. The variation in placebo types may indicate the different drug delivery route as placebo can’t be different from that. Each of the topic (keyword) present in the map and their co-occurrence highlight the different aspects of the clinical trials. Umbilical Cord Mesenchymal Stem Cells (UC MSC), for example is also now used to reduce mortality and morbidity among severe COVID-19 patients. [12] Keeping in view the plausible adverse impact of the repurpose drugs, they are being administered along with following standard of care (standard care) protocol. This explains the occurrence of standard care in this map. The map thus speaks to a large audience.

Looking more closely at the map, we find repurposed drugs as well as new trials primarily belong to the category of ‘antiviral drug’. Antiviral drugs were also the first repurposed drugs against SARS-CoV-2 infection. Antivirals targets the virus’ life cycle at stages like host cell binding, uncoating, synthesis and translation of viral mRNA, “release of newly synthesised virus” etc. [13] Due to these activities of the antivirals they can be used for prophylaxis, virus suppression, preventing severe outcomes in patients etc. Hydroxychloroquine, an antimalarial drug, has shown to have antiviral activity against SARS-CoV-2.

One can trace the key studies that may have motivated the clinical trial. The strong linkage seen between hydroxychloroquine and Azithromycin is a result of ongoing clinical trial using this drug combination. This can be traced to the study by Gautret et al. [14] that the combination of these two drugs “was significantly more efficient for virus elimination”. Similarly research studies also have led to a drug being dropped or provide a caution to ongoing trail. This can be seen in case of hydroxychloroquine.

Linkages seen in the map such as between Ivermectin-Chloroquine, Ivermectin-Hydroxychloroquine point out that clinical trials of these drugs are being done together in a synergistic manner. Senanayake [15] offered a “perspective that antiviral combinations with a ‘double hit effect’ may offer the best chance of success and clinical translatability”. Ivermectin, an antiparasitic drug, is shown by Caly et al. [16] to reduce viral replication. On the other hand Hydroxychloroquine according to Patri and Fabbrocini [17] could “interfere with the glycosylation of angiotensin-converting enzyme 2” which can reduce the “binding efficiency” of SARS-CoV-2 spike protein and ACE2 of host cells. Hence, their combination “could act in consequential and synergistic manner”. [17]

A closer look at why linkages are observed between Lopinavir-Ritonavir and Remdesivir, Hydroxychloroquine-Remdesivir require a deeper analysis. These types of combinations are happening because some clinical trials are using various arms in a single trial. In other words these trials are using single or various drug combinations in a clinical trial. Solidarity trails are also using this strategy.

The National Institute of Health (NIH) has recommended Remdesivir in “SpO2 ≤94% on ambient air (at sea level)” and for patients who are on mechanical ventilation. The drug however according to NIH has insufficient data backing to recommend it for mild to moderate patients. Many trials for example of Asan Medical Center (Trial ID NCT04307693) has three arms: Lopinavir/Ritonavir (experimental arm), Hydroxychloroquine (Active comparator arm), and Control arm (no intervention). This phase 2 trial divides each arm in 1:1:1 ratio and tries to compare the effectiveness of these drugs to reduce viral load. Several other trials are also using arms of 5-6 drugs in their studies, therefore the linkages pattern between these drugs is seen. .It was found that SARS-CoV-2 interferes with the blood pressure maintaining hormone Angiotensin II leading to high blood pressure and lung damage.[18] Losartan, a blood pressure medication and part of the dark blue cluster, is being tested to block the hormone thereby preventing lung damage. It is observed in patients that the inflammation triggering protein Interleukin-6 (IL-6) (cytokine) is overexpressed resulting in cytokine storms. This can lead to hyper-inflammation thus causing damage to lung and other organs which can be life threatening. Therefore, Tocilizumab which can “block cellular receptors for interleukin-6 (IL-6)” [19] is being used in the clinical trials to prevent cytokine storms in patients.

Thus, the maps bring out some salient characteristics of ongoing clinical trials and provides important signals.

## Discussion and Conclusion

A very intensive research activity and clinical trials can be observed surrounding COVID-19. The present paper attempts to capture salient aspects of the ongoing research and clinical trials drawing from the huge volume of ongoing research papers and clinical trials. Using sophisticated analytical tools the research trend, influential papers, and map of ongoing clinical trials that dominate the overall landscape of clinical trials were identified. Informed assessment was made by undertaking content analysis of the influential papers and exploring the rationale behind the linkages observed and their implications.

The trends of global research and clinical trials shows the speed at which extensive research and clinical trials is undertaken to understand the various aspects of this disease and finding an effective treatment for the disease. The ten most influential papers identified based on their altmetric score show that they have actively been covered in news and policy documents. Another common characteristic of these papers are they have been published in *highly reputed journal* which plausibly was also a factor in attracting high online influence. Other indicators highlight the dissemination of these papers in the diverse community from public, practitioners to scientists. This type of fast and diverse dissemination of research papers underscores the importance of this research for academics, practitioners and society. Content analysis of these ten influential papers draw attention to the importance of these papers in identifying virus characteristic, drug treatment/treatment protocols, disease contamination, epidemic model, etc. Examining cited policy documents and clinical studies shows their wide reaching impact.

The map reasserts the importance of repositioning as a key strategy undertaken for addressing this disease as numbers of repurposed drugs were identified in the clinical trials map. The importance of the map can be seen in terms of giving key signals that it provides. Dexamethasone for example was identified in the map based on the number of trails and linkages. In other words it was identified as an important drug in clinical trial. This repurposed drug has been shown effective in reducing mortality based on the extensive clinical study result of Oxford University. The map projected in this study had identified this as one of the key drugs in the clinical trials.

The study based on application of advanced information retrieval tools and using sophisticated visualisation methods has drawn attention to some salient aspects of research and clinical trials being undertaken. The importance of this approach can be appreciated if one looks at it in the perspective of the huge volume of research papers and clinical trials from which the top ten papers and map of clinical trials was created. The content analysis of papers and understanding the linkage in the clinical map with ongoing studies has helped in drawing deeper insights. This paper thus provides a useful approach to capture new research and innovation insights and goes beyond quantitative analysis that are undertaken primarily of research papers. A more informed view, however, can only be possible if further qualitative analysis is undertaken.

## Data Availability

Data extracted from Dimensions database

